# Clonal hematopoiesis is enriched in melanoma and associated with genotype-specific differences in tumor growth and survival

**DOI:** 10.64898/2026.07.13.26357981

**Authors:** Mason Alford-Holloway, Sarah C. Reed, Yash Pershad, Joseph C. Van Amburg, Chad Potts, Sanjay R. Mohan, Leo Y. Luo, P. Brent Ferrell, Michael R. Savona, Ben Ho Park, Douglas B. Johnson, Alexander G. Bick, Ashwin Kishtagari

## Abstract

**Background:** The clinical significance of clonal hematopoiesis of indeterminate potential (CHIP) in melanoma remains incompletely defined, particularly with respect to CHIP genotype, clone size, and somatic mutations (e.g *BRAF* mutations). We integrated human cohort data and a syngeneic melanoma mouse model to evaluate whether CHIP is associated with melanoma risk, tumor growth, and differential clinical outcomes.

**Methods:** We analyzed CHIP prevalence and survival in a large treatment-unselected melanoma cohort (n=2,480), evaluated tumor growth in a syngeneic *BRAF*-mutant (*BRAF*^mut^) melanoma murine model of *TET2*-CHIP and *DNMT3A*-CHIP, and assessed survival outcomes in an immune checkpoint inhibitor (ICI)-treated advanced melanoma cohort (n=361). Associations with progression-free survival (PFS) and overall survival (OS) were evaluated using Kaplan-Meier analyses and multivariable Cox proportional hazards models.

**Results:** CHIP was enriched among patients with treatment-unselected melanoma compared with age/sex-matched healthy controls, and larger CHIP clone size showed an age-adjusted association with inferior OS. In a syngeneic *BRAF*^mut^ melanoma murine model, *TET2*-CHIP, but not *DNMT3A*-CHIP, was associated with significantly increased primary melanoma tumor growth. Among patients with ICI-treated advanced melanoma, CHIP was associated with worse OS compared with patients without CHIP. *TET2*-CHIP had the strongest adverse association with survival, whereas *DNMT3A*-CHIP was not significantly associated with PFS or OS.

**Conclusions:** CHIP is enriched in melanoma and exploratory analyses demonstrate genotype-specific differences in melanoma tumor growth and clinical outcomes. These findings support further investigation of genotype-specific CHIP profiling as a potential biomarker for melanoma risk stratification and immunotherapy outcomes.

**What is already known on this topic:** Clonal hematopoiesis of indeterminate potential (CHIP) is an age-associated condition linked to inflammation, cardiovascular disease, hematologic malignancy, and adverse outcomes across several cancer settings. Prior studies have suggested that CHIP may influence immune function and response to immune checkpoint inhibitors, but the clinical relevance of CHIP in melanoma, particularly by CHIP genotype, clone size, and tumor *BRAF* genotype, remains incompletely defined.

**What this study adds:** This study shows that CHIP is enriched among patients with melanoma and that larger CHIP clone size is associated with inferior overall survival in age-adjusted analyses. In complementary murine and clinical cohorts, *TET2*-CHIP, but not *DNMT3A*-CHIP, was associated with increased melanoma tumor growth and worse survival outcomes among patients with ICI-treated advanced melanoma, with exploratory evidence that CHIP-associated risk may be most pronounced in *BRAF*-mutant melanoma.

**How this study might affect research, practice or policy:** These findings support further investigation of CHIP as a genotype-specific biomarker of melanoma biology that has the potential to refine risk stratification for patients with advanced melanoma. They further motivate mechanistic studies of how hematopoietic mutations shape antitumor immunity in specific tumor genomic contexts.

## BACKGROUND

Clonal hematopoiesis of indeterminate potential (CHIP) is an age-related [1–3] phenomenon in which leukemia-associated somatic mutations, most commonly in the epigenetic regulator genes *DNMT3A*, *TET2*, and *ASXL1*, occur in hematopoietic stem and progenitor cells and promote clonal expansion of blood cells [4–5]. Current guidelines identify CHIP in patients if the mutation is present in at least 4% of blood cells (2% variant allele frequency) and another clonal hematologic disorder is not present [6]. Individuals with CHIP have increased rates of hematologic malignancy, cardiovascular disease, and worse all-cause mortality compared to individuals without CHIP [7–9]. Additional studies have highlighted that CHIP is biologically heterogeneous: different CHIP mutations endow distinct clonal growth kinetics [10] and are differentially associated with a variety of disease outcomes and signaling pathways [11–14]. While initially described for its association with myeloid neoplasms, CHIP is now well-recognized as a systemic modifier of inflammation, thrombosis, and immunity [15–16], with broad implications for patients with solid tumors who are exposed to cytotoxic, targeted, and immune therapies.

The clinical significance of CHIP in the biology and evolution of solid tumors is an active area of research. Recent studies have demonstrated that patients with solid tumors have high rates of CHIP [17–18], and many cancer therapies further promote expansion of CHIP clones by directly altering the fitness landscape of clonal hematopoiesis [19–21]. Several mechanistic studies have demonstrated that perturbations in CHIP-driver genes (e.g., *TET2* and *DNMT3A*) can reprogram lymphocyte function [22–23] and lead to increased expression of inflammatory genes in innate immune cells [24–28], suggesting a potential mechanism through which CHIP might alter tumor immunology, impact response to cancer therapy, and influence clinical outcomes in solid tumors. Specifically, in the context of melanoma, recent analyses have demonstrated that *TET2* and *DNMT3A*-driven CHIP clones expand following initiation of immune checkpoint inhibitor therapy [29]. CHIP has also been shown to be associated with worse OS after ICI treatment across multiple solid tumor types, including melanoma [30]. Recent translational studies further underscore the complexity of the relationship between TET2-CHIP and ICI response, including evidence that hematopoietic TET2 inactivation can enhance response to immune checkpoint blockade in murine tumor models by reprogramming the myeloid compartment toward a more immunostimulatory state, thereby promoting phagocyte-mediated antigen presentation and supporting CD4+ and CD8+ T-cell infiltration and effector function within the tumor microenvironment. [31]. Similarly, *TET2*-CHIP has been shown to promote an antigen-presenting macrophage state, increase intratumoral T-cell infiltration, and improve checkpoint therapy response in solid tumor models [32]. Together, these studies suggest that *TET2*-CHIP can reshape the tumor-immune microenvironment in ways that alter ICI responsiveness, but that the direction and magnitude of this effect may depend on clone size, hematopoietic composition, and the pre-existing inflammatory state of the tumor microenvironment. Thus, rather than functioning as a uniform biomarker of ICI resistance or sensitivity, CHIP may act as a genotype- and context-dependent modifier of antitumor immunity.

Melanoma is a particularly compelling context in which to investigate CHIP. Patients with advanced melanoma are often in the age range where CHIP is common, and are increasingly treated with ICIs in the first-line setting regardless of *BRAF* status [33]. In preclinical models, *TET2* deficiency in tumor-infiltrating myeloid cells enhances immunosuppressive function and promotes melanoma progression, whereas *TET2* activity in tumor cells and immune cells can govern chemokine production, PD-L1 expression, and lymphocyte infiltration [34–35]. At the same time, *BRAF*^V600E^ mutations, present in roughly 40-50% of cutaneous melanomas, shape both tumor biology and treatment pathways [36]. Targeted *BRAF*/*MEK* inhibition induces rapid tumor regressions but is frequently limited by acquired resistance, whereas ICIs can produce more durable remissions [37–39]. Whether CHIP, particularly *TET2*- or *DNMT3A*-driven clones, interacts with tumor *BRAF* mutation status to influence melanoma progression or ICI benefit has not been systematically explored.

## METHODS

### BioVU melanoma cohort

We conducted a cohort study using BioVU, the Vanderbilt University Medical Center (VUMC) biorepository linking electronic health records to whole-genome sequencing data from 242,871 participants from 2006 to 2025 [40]. Of these participants, 2,480 patients had a melanoma diagnosis across all stages and without selecting for prior treatment history. To evaluate enrichment of CHIP among melanoma cases, we compared individuals with melanoma against three control definitions: cancer-free control (n=206,519), all non-melanoma control (n=240,391), and 1:4 age/sex-matched control adjusted for age, sex, and 10 ancestry principal components (n=9,920). Adjusted odds ratios and 95% confidence intervals were estimated by exponentiating the CHIP regression coefficient. Within melanoma cases with available somatic sequencing, we evaluated CHIP prevalence in *BRAF*^mut^ versus *BRAF*^wt^ melanoma. For survival analyses in BioVU melanoma cases, OS was defined from melanoma diagnosis to death from any cause; ICI exposure and stage IV disease were included as covariates where applicable because the BioVU cohort was not selected on the basis of ICI treatment. Because several *BRAF*-mutant melanoma cases were sparse, OS models were additionally fit with Firth penalization; we report Firth (age-adjusted) hazard ratios and flag cells where standard models failed to converge.

### Syngeneic *BRAF*-mutant melanoma model

All animal procedures were approved by the Institutional Animal Care and Use Committee (IACUC) and performed according to VUMC and IACUC regulatory guidelines. Mice with *Tet2*-CHIP (n=8) and *Dnmt3a*-CHIP (n=8) were generated as previously described [41]. Briefly, recipient male mice were sub-lethally irradiated and then injected with a mixture of bone marrow harvested from wild-type and *Dnmt3a*-mutant (*Dnmt3a* R878H/+) or *Tet2*-mutant (*Tet2*-/-) donor mice. Control mice received wild-type floxed cells lacking Cre recombinase rather than mutant cells but were otherwise identical.

After 12 weeks of engraftment, the resulting chimeric mice were injected subcutaneously with the syngeneic melanoma line Rich 1.1 [42]. Tumors were grown for up to six weeks, with endpoint defined as tumor ulceration of any mouse in the cohort. Mice were euthanized at endpoint, and tumors were weighed. Differences in tumor weight between CHIP and control mice were statistically evaluated using a two-sample t-test with alpha = 0.05. Experimenters who injected melanoma cells and measured tumors were blinded to mouse genotype until after all data were collected.

### ICI-treated advanced melanoma clinical cohort

This single-institution, retrospective cohort study was approved by the Institutional Review Board at Vanderbilt University Medical Center (VUMC) (IRB #100178). The ICI-treated melanoma cohort included 361 participants with unresectable stage III/IV melanoma who underwent whole blood-based genomic sequencing between 2014 and 2024 prior to initiation of anti-PD-1-based ICI therapy. Only patients with a diagnosis of stage III/IV unresectable melanoma were included. All clinicopathologic characteristics and clinical outcomes were gathered by manual chart review. A cohort of 6,498 healthy age/sex-matched participants were used to estimate background CHIP prevalence. CHIP prevalence was then compared between the ICI-treated melanoma cohort and the 1:18 age/sex-matched healthy cohort using a chi-square test.

### Detection of CHIP and tumor BRAF mutation calling

Whole blood samples obtained prior to initiation of ICI therapy were used for CHIP calling as previously described [43]. Briefly, we performed error-corrected targeted sequencing of the exons of CHIP-driver genes with median depth after deduplication of approximately 1700x. To detect CHIP-associated variants, Mutect2 was used to call somatic mutations in CHIP-driver regions. We filtered variants based on read depth (≥100 for deep sequencing, ≥15 for genome sequencing), variant allele read depth (≥3 for deep sequencing, ≥2 for genome sequencing), double-strand support, inconsistency with germline heterozygosity, and VAF ≥0.5% [44]. For primary analyses, CHIP was defined as a somatic mutation in a CHIP-driver gene with VAF ≥2%. Variants with VAF <2% were considered subthreshold CHIP-associated variants and were classified as CHIP-negative for primary analyses. Genome sequencing was performed using the Illumina DNA PCR-Free Prep targeting a median depth of 30x when available. Tumor *BRAF* genotype was determined by tumor next-generation sequencing or other clinically available assays used as standard of care, including PCR-based testing when applicable.

### Statistical Analysis

Baseline clinicopathologic characteristics were summarized by CHIP status and compared using Fisher’s exact tests for categorical variables and Mann-Whitney U tests for continuous variables. For the large, treatment-unselected BioVU melanoma cohort, overall survival was defined as the interval from melanoma diagnosis to death from any cause. For the ICI-treated advanced melanoma cohort, PFS was defined as the interval from initiation of anti-PD-1–based therapy to disease progression or death, and OS was defined as the interval from therapy initiation to death from any cause. Patients without an event were censored at the date of the last follow-up. Survival distributions were visualized using the Kaplan–Meier method and compared using the log-rank test.

For primary survival analyses for the ICI-treated melanoma cohort, Cox proportional hazards regression was used to estimate hazard ratios and 95% confidence intervals (CIs). Models evaluating the association of CHIP with PFS or OS compared to patients without CHIP were adjusted for age, sex, *BRAF* mutation status, stage at anti-PD-1 initiation, and prior therapy. Models evaluating the association between clone size and outcomes using maximum VAF categories overall and within *TET2*-CHIP and *DNMT3A*-CHIP subsets, again using patients without CHIP as the reference group, were similarly adjusted for age, sex, *BRAF* mutation status, stage at anti-PD-1 initiation, and prior therapy before anti-PD-1.

To evaluate whether the association between CHIP and survival differed by tumor *BRAF* genotype for the ICI-treated cohort, we performed *BRAF*-stratified Cox proportional hazards analyses. Separate models were fit within the *BRAF*-mutant and *BRAF*-wild-type parent groups, with the corresponding no-CHIP subgroup within each *BRAF* stratum serving as the reference.

To formally evaluate effect modification by tumor *BRAF* genotype, we fit Cox proportional hazards models including main effects for CHIP status and *BRAF*-mutant status, as well as a CHIP × *BRAF* interaction term. Interaction was evaluated separately for PFS and OS using Wald tests for the interaction coefficient and likelihood ratio tests comparing models with and without the interaction term. Because subgroup sizes were limited, especially for gene-specific CHIP analyses, interaction analyses were interpreted as exploratory. Additive interaction was evaluated using the relative excess risk due to interaction (RERI), estimated from models parameterized with a four-level CHIP/BRAF exposure variable, with confidence intervals estimated by bootstrap resampling.

We also fit multivariable Cox proportional hazards models in the full ICI-treated cohort to evaluate the independent associations of tumor- and CHIP-related covariates with PFS and OS. Covariates included *BRAF*-mutant status, any CHIP, CHIP with maximum VAF ≥10%, combined CHIP and *BRAF*-mutant positivity, age >65 years, stage IV disease at anti-PD-1 initiation, presence of ≥2 CHIP clones with VAF ≥2%, and baseline LDH ≥225 U/L. Adjusted HRs with 95% CIs were reported, and results were displayed as tabular forest plots on a logarithmic scale with HR=1 as the null reference. This joint-exposure term was interpreted descriptively and was not used as the formal test of interaction. P-values for Cox models were obtained using Wald tests.

All statistical tests were two-sided, and p < 0.05 was considered statistically significant. Analyses were performed in R (v4.x), primarily using the survival and survminer packages. GraphPad Prism (v10.3.1) was used for analysis and plotting of murine tumor data.

## RESULTS

### CHIP is enriched among treatment-unselected melanoma cases

In the combined BioVU melanoma case and cancer-free control population, CHIP was identified in 10,592 of 208,999 individuals (**Table 1**). Individuals with CHIP were substantially older than those without CHIP, and the CHIP-positive group had a slightly higher proportion of male individuals compared with the non-CHIP group. Race distributions differed by CHIP status, with a higher proportion of White individuals among those with CHIP (81.9% vs 77.4%, p < 0.001) and lower proportions of Black/African American individuals (10.5% vs 13.2%, p < 0.001) and Asian individuals (1.2% vs 1.9%, p < 0.001). Among individuals with CHIP, *DNMT3A* mutations were the most common CHIP-associated alteration, present in 4,859 individuals (45.9%), followed by *TET2* mutations in 3,044 individuals (28.7%). Melanoma cases were more frequent among individuals with CHIP than among those without CHIP (2.3% vs 1.1%, p < 0.001), supporting enrichment of CHIP among melanoma cases in this treatment-unselected institutional biorepository cohort.

**Table 1.**
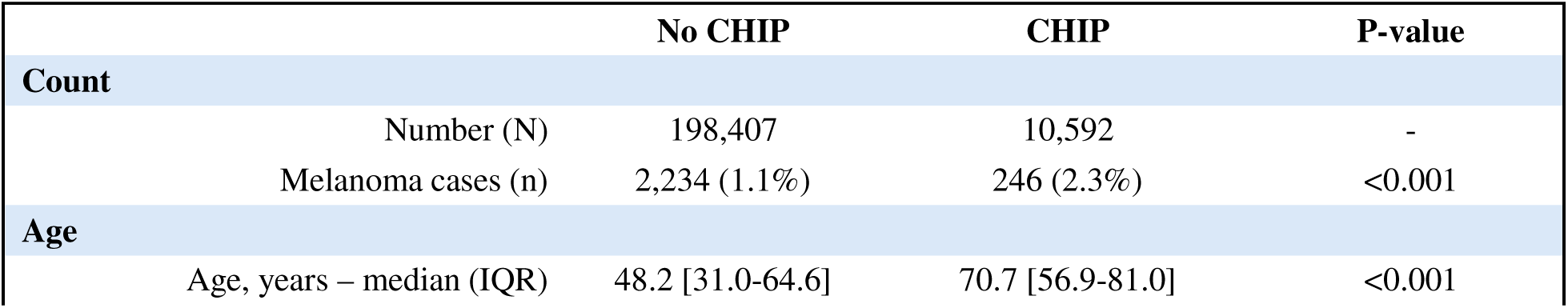
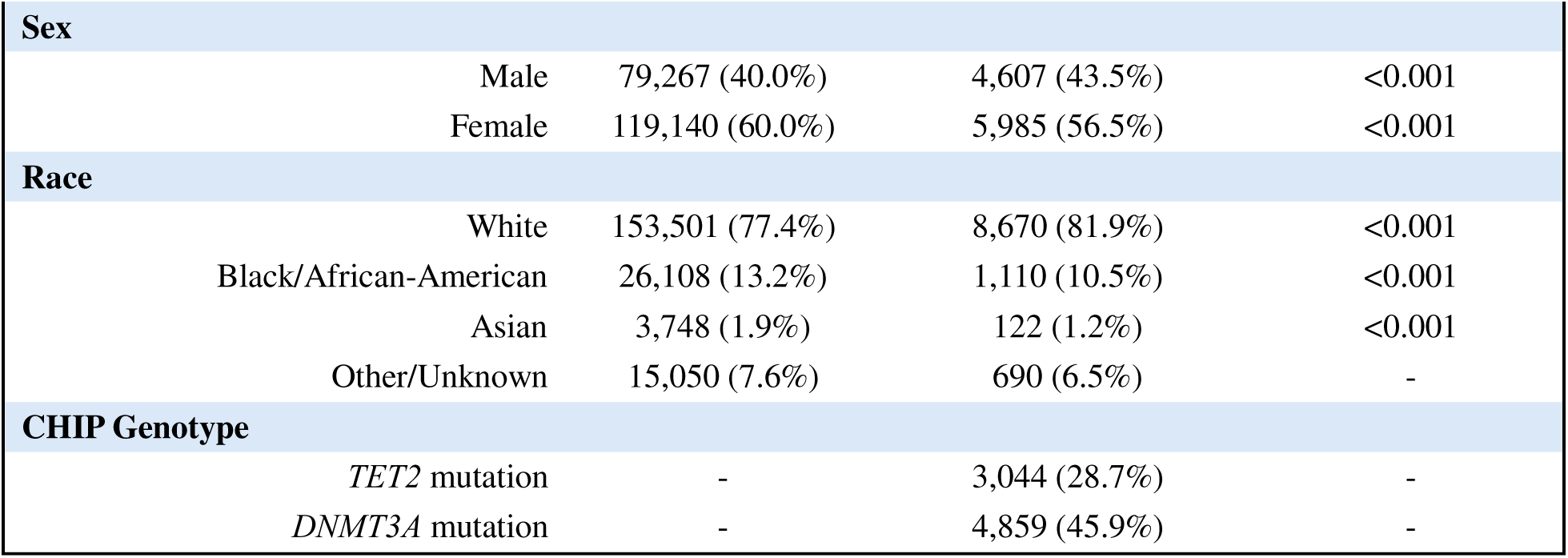
BioVU melanoma cases & cancer-free controls, by CHIP status.

In comparing the melanoma cases (n=2,480) within this institutional dataset versus controls, CHIP was significantly more common in melanoma across multiple control groups: OR was 1.61 (1.40–1.84) vs cancer-free controls, 1.50 (1.31–1.72) vs all non-melanoma controls, and 1.54 (1.32–1.80) in a 1:4 age/sex-matched set (**Supplementary Table 1**).

Among all-comer 2,480 melanoma cases, individuals with CHIP (n=246) were substantially older than those without CHIP (n=2,234), with a median age of 70.4 years versus 60.0 years, respectively (**Supplementary Table 2)**. There were no significant differences in sex or race distributions by CHIP status within the melanoma cohort. Among individuals with CHIP, *DNMT3A* mutations were the most common CHIP-associated alteration, present in 116 individuals (47.2%), followed by *TET2* mutations in 63 individuals (25.6%). A significantly larger proportion of melanoma patients without CHIP (11.6%) received ICI therapy compared to melanoma patients with CHIP (6.8%).

### CHIP prevalence among melanoma cases differs by tumor *BRAF* genotype

Within the institutional cohort, 647 cases had available somatic sequencing data regarding *BRAF* mutation status (**Supplementary Table 3**). Among these, 295 patients had *BRAF*^mut^ tumors and 352 patients had *BRAF*^wt^ tumors. Individuals with CHIP were substantially older than those without CHIP, with a median age of 65.6 years versus 56.9 years, respectively (p < 0.001). The CHIP-positive group had a similar proportion of male individuals compared with the non-CHIP group (62.5% vs 58.9%, p = 0.67). Race distributions did not significantly differ by CHIP status. Among individuals with CHIP and *BRAF* sequencing data, *DNMT3A* mutations were the most common CHIP-associated alteration, present in 25 individuals (44.6%), followed by *TET2* mutations in 16 individuals (28.6%).

Among patients within this treatment-unselected melanoma cohort with available *BRAF* somatic sequencing data, CHIP was less common in *BRAF*^mut^ tumors (adjusted OR 0.36, 95% CI 0.17– 0.72, p = 0.006), with *DNMT3A*-CHIP showing a concordant trend (OR 0.41, 0.14–1.07, p = 0.084) and *TET2*-CHIP non-significant and sparse (OR 0.70, p = 0.56) (**Supplementary Table 4**).

### Large CHIP clone size shows an age-adjusted association with inferior OS in treatment-unselected melanoma

Within the treatment-unselected institutional cohort CHIP at higher clone size (VAF ≥10%) showed a significant association with worse OS (age-adjusted HR 1.75, 95% CI 1.04–2.79, p = 0.035) that did not persist after adjustment for sex, ICI, and stage **(Figure 1)**. CHIP at VAF ≥2% was not significant (HR 1.35, 95% CI 0.81–2.15, p = 0.24), and gene-specific exposures (*TET2, DNMT3A*) were null at both thresholds. BRAF-stratified analysis limited by very small numbers of exposed deaths (1–7 per subgroup); though patients with *BRAF*^wt^ and CHIP showed a trend toward inferior survival (HR 2.16–2.39, p = 0.06–0.09, 7 deaths).

**Figure 1.**
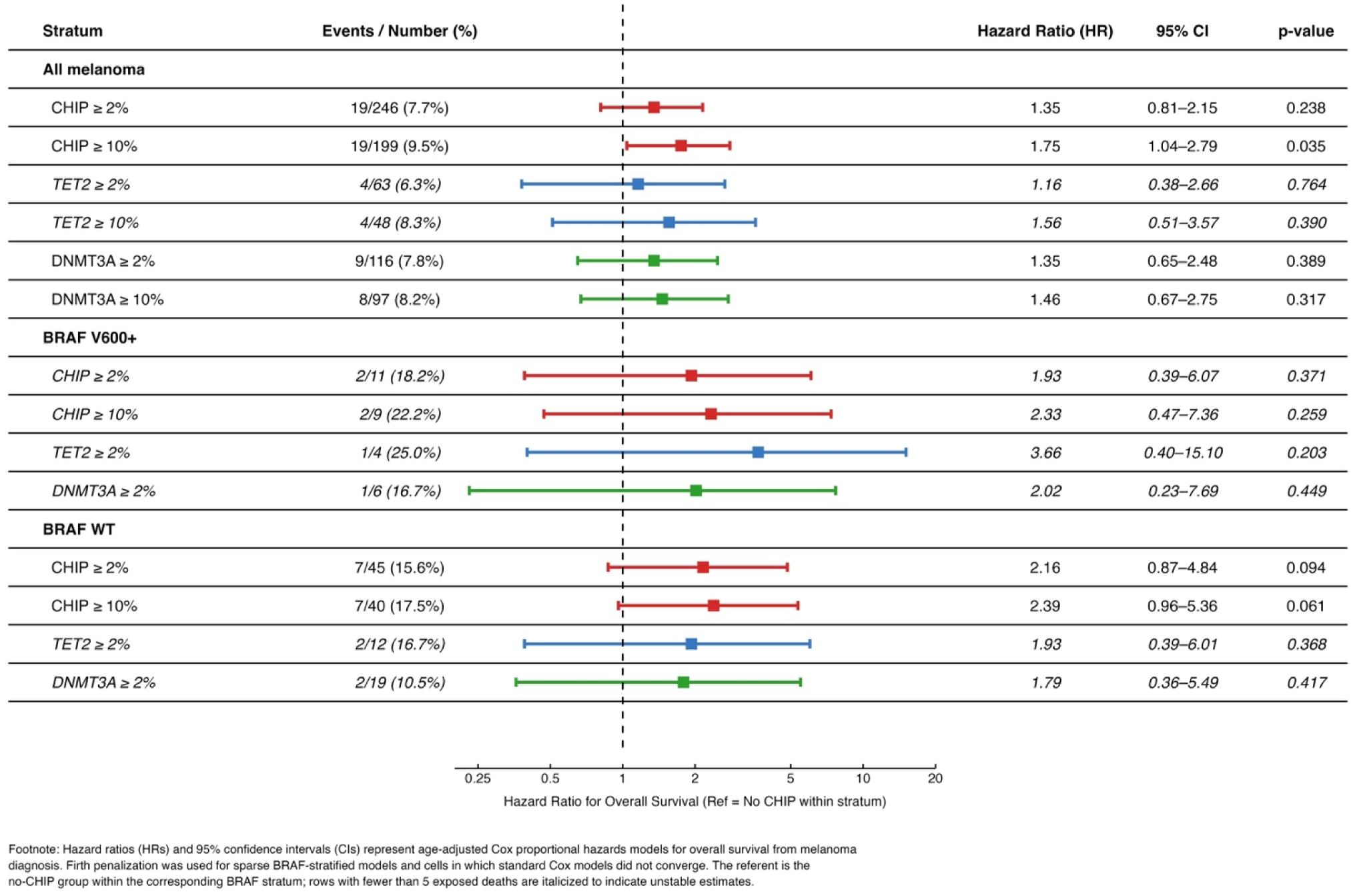
Tabular forest plot of Cox proportional hazards models evaluating the association of CHIP clone size and genotype with overall survival in the treatment-unselected melanoma cohort, stratified by *BRAF* genotype. The events/number column reports deaths within each subgroup. Squares represent adjusted hazard ratios (HRs) and horizontal whiskers denote 95% confidence intervals (CIs) on a logarithmic scale; the dashed vertical line indicates HR=1, with reference group defined as patients without CHIP within corresponding stratum. BRAF-stratified estimates were generated using Firth-penalized Cox models due to sparse events and non-convergence of standard Cox models in several cells. Rows with fewer than 5 exposed deaths are italicized to indicate unstable estimates.

### *TET2*-CHIP is associated with primary melanoma growth in a *BRAF*-mutant syngeneic mouse model

Given the observation that CHIP was enriched in melanoma patients compared to healthy controls in the institutional cohort, as well as previous studies demonstrating the genotype-specific impact of CHIP on solid tumor outcomes [41], we further explored this relationship in vivo. We performed chimeric bone marrow transplantation to generate mice with *DNMT3A*-CHIP (n=8) or *TET2*-CHIP (n=8), representing the two most commonly mutated CHIP genotypes. After transplants were engrafted, resulting in mice with a mixture of wild-type and CHIP-mutant hematopoietic stem cells, the syngeneic *BRAF*^mut^ melanoma cell line Rich1.1 was implanted subcutaneously (**Figure 2A**).

**Figure 2.**
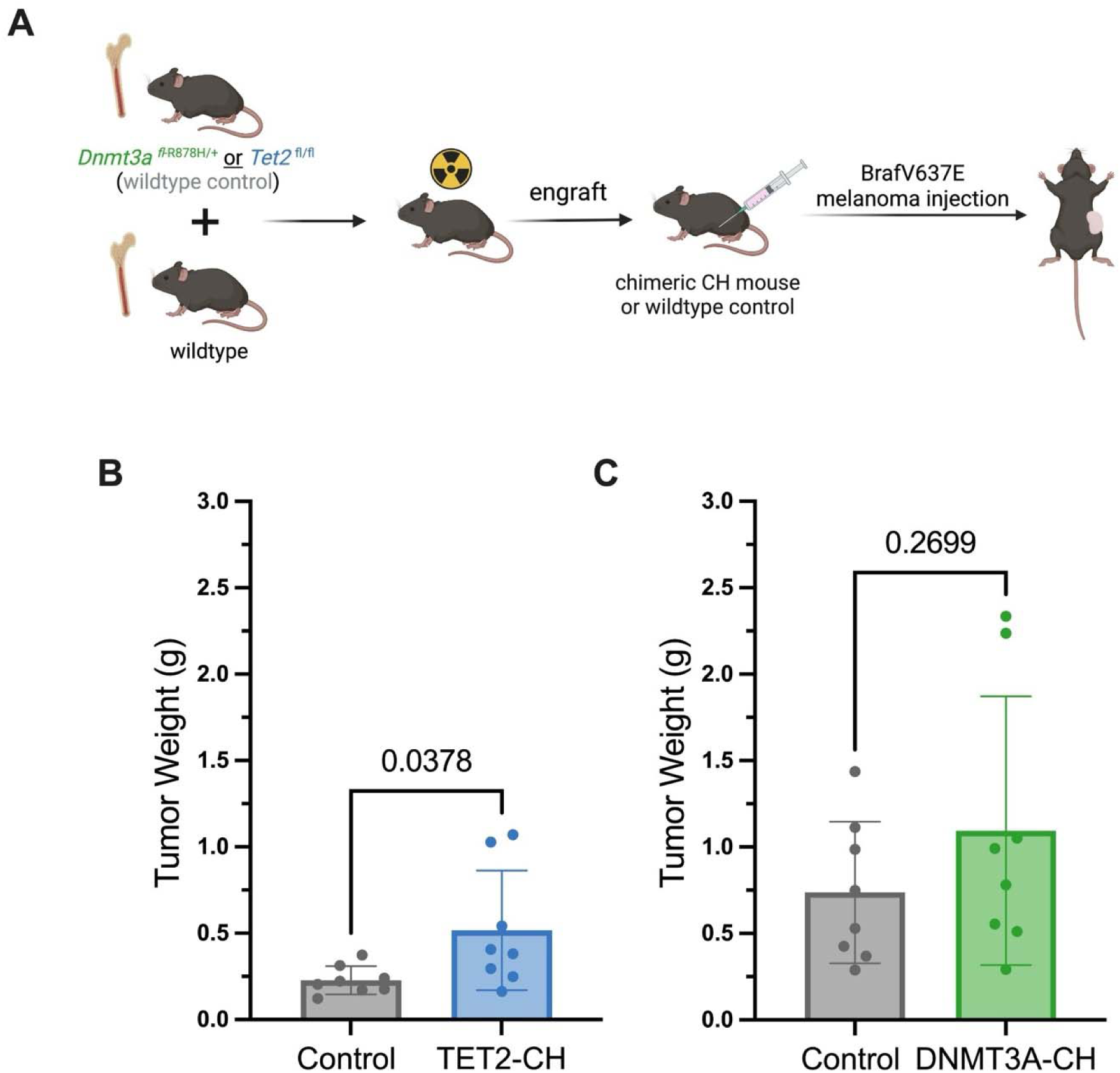
Primary melanoma tumor growth differs by CHIP genotype. **(A)** Bone marrow transplantation was used to generate chimeric mice with *TET2*-CHIP or *DNMT3A*-CHIP, which were subcutaneously injected with *BRAF*-mutant Rich1.1 melanoma cells after engraftment. **(B)** Tumor weight at endpoint (day 33) in *TET2*-CHIP mice and corresponding controls. **(C)** Tumor weight at endpoint (day 40) in *DNMT3A*-CHIP mice and corresponding controls. Differences in tumor weight were evaluated using t-tests.

Tumors in mice with *TET2*-CHIP were significantly larger than their controls (**Figure 2B**), while there was no difference in tumor size between mice with *DNMT3A*-CHIP and controls (**Figure 2C**). These data mirror our clinical observations and similar findings in other solid tumors, in which *TET2*-CHIP but not *DNMT3A*-CHIP results in increased primary tumor growth [41].

### CHIP is enriched in patients with ICI-treated advanced melanoma

Given the genotype-specific effect of CHIP on primary tumor growth observed in our *BRAF*^mut^ melanoma model, as well as previous studies demonstrating that the aberrant inflammatory milieu associated with CHIP modulates the efficacy of ICI therapy in solid tumor settings [31–32], we investigated the clinical significance of CHIP in a smaller, ICI-treated advanced melanoma discovery cohort. We identified 361 patients with unresectable stage III or stage IV melanoma who underwent whole blood-based genomic sequencing prior to initiation of ICI therapy between 2014 and 2024.

Among 361 melanoma patients who received ICI therapy, 100 patients (27.7%) had CHIP identified immediately prior to ICI initiation and 261 patients (72.3%) did not. There were no significant differences between the CHIP and no-CHIP subgroups in the distribution of baseline characteristics such as sex, stage at ICI initiation, or prior lines of treatment **(Table 2**). As expected, CHIP prevalence increased with age (**Supplementary Figure 1**). Among the melanoma patients with CHIP (n=100), 77 patients had a single CHIP clone, 19 patients had two distinct CHIP clones, and 4 patients had three or more distinct CHIP clones. Variants were detected in 12 CHIP-related genes, with *DNMT3A* (n=48, 48.0% of CHIP cohort) and *TET2* (n=32, 32% of CHIP cohort) representing the most common driver mutations (**Supplementary Figure 2**). Compared with an age- and sex-matched healthy control cohort, CHIP prevalence was significantly higher among melanoma patients (**Supplementary Figure 1**).

**Table 2.** Baseline characteristics of ICI-treated advanced melanoma cohort.

|  | No CHIP | CHIP | P-value |
| --- | --- | --- | --- |
| Age |  |  |  |
| Mean, years (SD) | 66.1 (9.6) | 71.4 (9.8) | <0.001 |
| Median, years [Min, Max] | 65.0 [46, 91] | 71.0 [47, 95] | <0.001 |
| Sex |  |  | 0.3228 |
| Male | 175 (67.0%) | 61 (61.0%) |  |
| Female | 86 (33.0%) | 39 (39.0%) |  |
| Stage at anti-PD-1 Start |  |  | 0.0894 |
| III | 17 (6.5%) | 4 (4.0%) |  |
| M1a | 50 (19.2%) | 25 (25.0%) |  |
| M1b | 47 (18.0%) | 15 (15.0%) |  |
| M1c | 105 (40.2%) | 49 (49.0%) |  |
| M1d | 42 (16.1%) | 7 (7.0%) |  |
| Prior Treatment |  |  | 0.1256 |
| Chemotherapy | 4 (1.5%) | 4 (4.0%) | 0.2245 |
| Radiation | 39 (14.9%) | 12 (12.0%) | 0.6126 |
| BRAF/MEK inhibitor | 32 (12.3%) | 7 (7.0%) | 0.1857 |
| Surgery | 27 (10.3%) | 15 (15.0%) | 0.2703 |
| Immunotherapy (Ipilimumab) | 55 (21.1%) | 17 (17.0%) | 0.4623 |
| Immunotherapy (Pembrolizumab) | 4 (1.5%) | 3 (3.0%) | 0.4014 |
| Immunotherapy (Nivolumab) | 6 (2.3%) | 0 (0.0%) | 0.1927 |
| Baseline Labs |  |  |  |
| LDH (mean) | 291.9 | 376.4 | 0.1717 |

### *TET2*-CHIP is associated with inferior survival in ICI-treated advanced melanoma

ICI-treated melanoma patients with CHIP demonstrated significantly worse OS compared with patients without CHIP (adjusted HR 1.43, 95% CI 1.05-1.95, p=0.023) and a trend toward worse PFS (adjusted HR 1.31, 95% CI 0.98-1.75, p=0.071) (**Figure 3A-B**). To evaluate whether these associations differed by CHIP genotype, we compared patients with *TET2*-CHIP (n=32) and *DNMT3A*-CHIP (n=48) with patients without CHIP (n=261). Patients with *TET2*-CHIP demonstrated significantly worse OS (adjusted HR 1.64, 95% CI 1.01-2.64, p=0.044) and a trend toward worse PFS (adjusted HR 1.55, 95% CI 0.98-2.45, p=0.060) (**Figure 3C-D**). In contrast, *DNMT3A*-CHIP was not significantly associated with either OS (adjusted HR 1.42, 95% CI 0.95-2.11, p=0.084) or PFS (adjusted HR 1.36, 95% CI 0.93-1.98, p=0.111) (**Figure 3E-F**). These findings suggest that the adverse association of CHIP with outcomes in melanoma is not uniform across genotypes and is most apparent among patients with *TET2*-CHIP.

**Figure 3.**
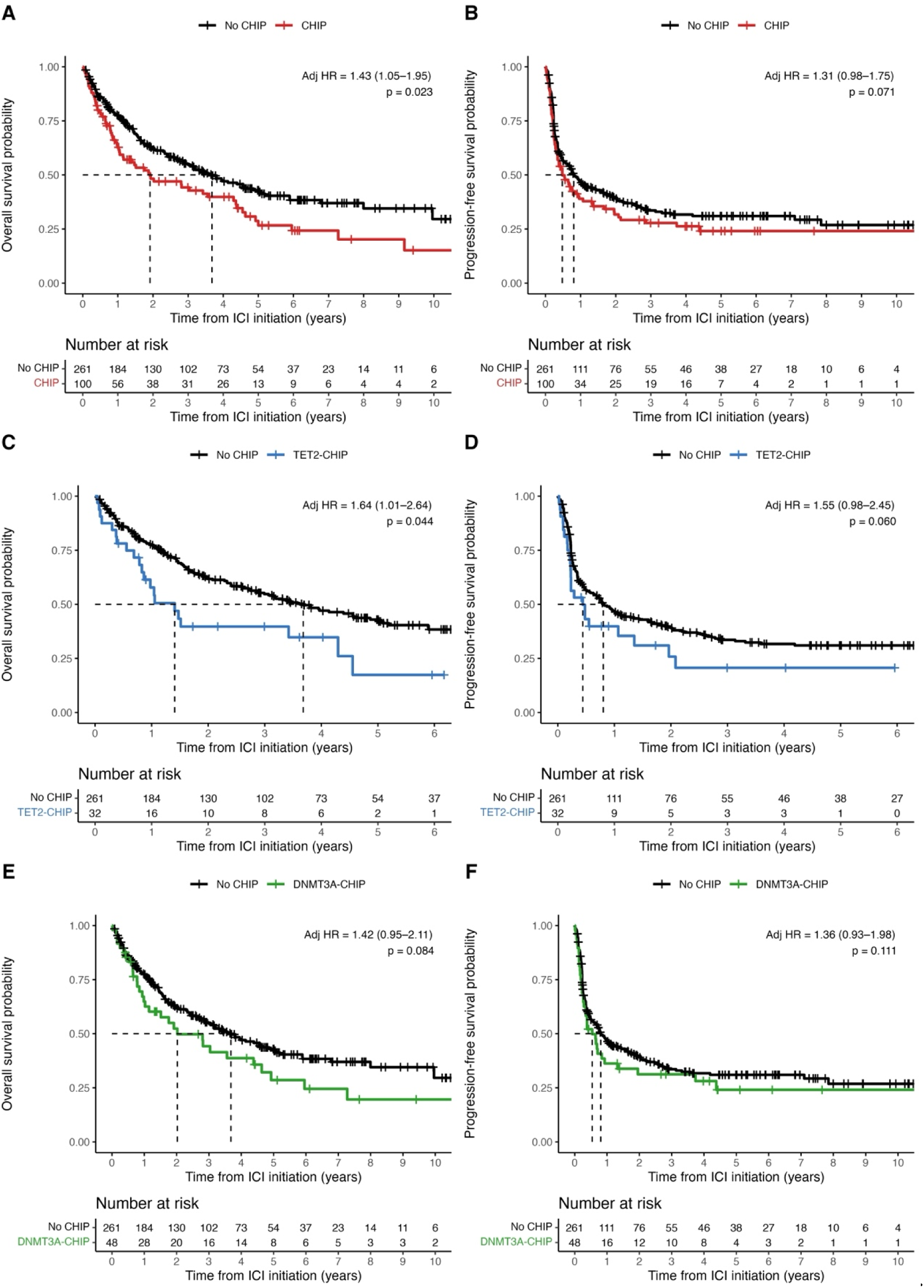
Kaplan-Meier curves comparing survival outcomes between melanoma patients with and without CHIP, and between gene-specific CHIP subgroups and patients without CHIP. **(A)** OS for patients with CHIP versus no CHIP. **(B)** PFS for patients with CHIP versus no CHIP. **(C)** OS for patients with *TET2*-CHIP versus no CHIP. **(D)** PFS for patients with *TET2*-CHIP versus no CHIP. **(E)** OS for patients with *DNMT3A*-CHIP versus no CHIP. **(F)** PFS for patients with *DNMT3A*-CHIP versus no CHIP. Hazard ratios and 95% confidence intervals are from Cox proportional hazards models adjusted for age, sex, *BRAF* genotype, stage at anti-PD-1 initiation, and prior therapy before anti-PD-1.

### Evaluating the clinical significance of CHIP clone size in ICI-treated melanoma

To evaluate the impact of CHIP clone size on clinical outcomes, we stratified patients according to maximum CHIP VAF and compared each group to patients without CHIP (**Figure 4**). Micro-CH was included in these analyses in recognition of the increasingly common detection of sub-2% VAF clones in patient samples due to deep sequencing [45]. Patients with micro-CH (VAF <2%) did not differ significantly from the no-CH reference for either PFS or OS, whereas patients with CH at VAF ≥2% demonstrated a trend toward worse PFS and significantly worse OS (HR 1.45, 95% CI 1.04-2.03, p=0.03). Patients with VAF ≥10% did not significantly differ from the no-CHIP group for either endpoint, likely reflecting limited sample size. These VAF non-mutually exclusive threshold categories for VAF ≥2% and VAF ≥10%, consistent with the analytic approach used for the corresponding figures.

**Figure 4.**
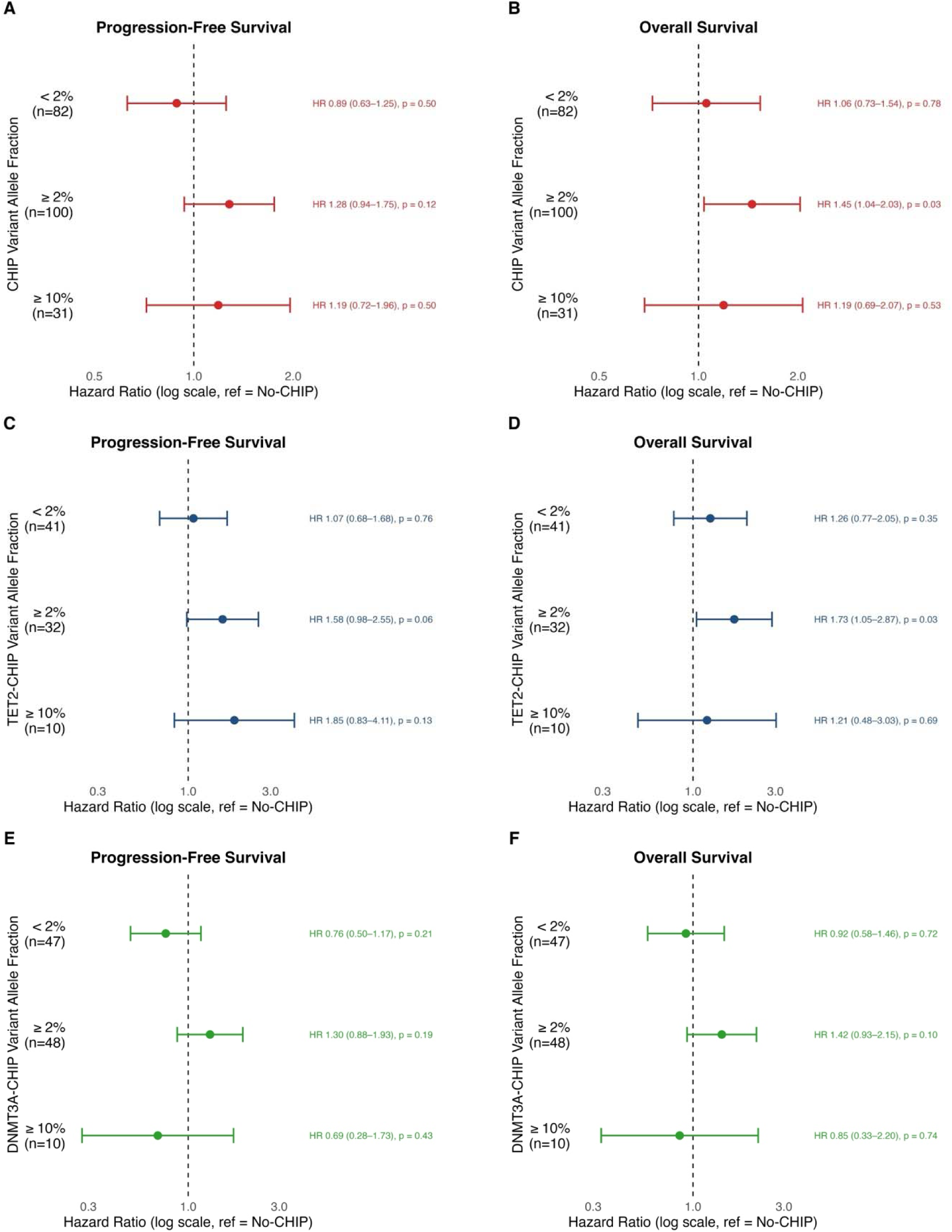
Association between CHIP clone size and survival outcomes. **(A)** PFS according to maximum CHIP VAF threshold category (<2%, ≥2%, and ≥10%). **(B)** OS according to maximum CHIP VAF threshold category (<2%, ≥2%, and ≥10%). **(C)** PFS according to *TET2*-CHIP VAF threshold category. **(D)** OS according to *TET2*-CHIP VAF threshold category. **(E)** PFS according to *DNMT3A*-CHIP VAF threshold category. **(F)** OS according to *DNMT3A*-CHIP VAF threshold category. For all comparisons, patients without CHIP served as the reference group.

When analyses were restricted to *TET2*-CHIP, genotype-specific differences became more apparent. *TET2* micro-CH was not associated with significantly different outcomes compared with no CH. In contrast, *TET2*-CHIP at VAF ≥2% was associated with a trend toward worse PFS and significantly worse OS. *TET2*-CHIP at VAF ≥10% was not significantly associated with either PFS or OS, again in the setting of small numbers.

By contrast, *DNMT3A*-CHIP was not significantly associated with PFS or OS across any VAF stratum. Together, these findings indicate that the prognostic impact of CHIP in melanoma is both genotype-specific and VAF-dependent, with the clearest adverse signal observed in *TET2*-CHIP, while *DNMT3A*-CHIP remained unassociated with outcomes even after stratification by clone size.

### Exploratory BRAF-stratified analyses of CHIP-associated risk

Because *BRAF* status is an important determinant of melanoma biology, we next evaluated the clinical significance of tumor *BRAF* genotype within our ICI-treated advanced melanoma cohort. Patients with *BRAF*^mut^ melanoma demonstrated significantly shorter PFS than patients with *BRAF*^wt^ melanoma (adjusted HR 1.51, 95% CI 1.12-2.03, p=0.006), whereas OS did not differ between the two groups (adjusted HR 0.99, 95% CI 0.70-1.39, p=0.951) (**Supplementary Figure 3**).

Given that tumor *BRAF* status was associated with differential outcomes in our cohort, we then evaluated whether the association between CHIP and survival differed by *BRAF* genotype (**Figure 5**). Among *BRAF*^mut^ patients, CHIP was not significantly associated with PFS (HR 1.59, 95% CI 0.84-3.01, p=0.156) but was associated with significantly worse OS (HR 2.37, 95% CI 1.17-4.80, p=0.017). Further, *TET2*-CHIP showed the strongest adverse association with OS in *BRAF*^mut^ patients (HR 7.85, 95% CI 2.07-29.74, p=0.002), although the number of *TET2*-CHIP cases was small (n=3). *DNMT3A*-CHIP in *BRAF*^mut^ disease was not significantly associated with either PFS (HR 1.72, 95% CI 0.81-3.66, p=0.160) or OS (HR 2.08, 95% CI 0.84-5.12, p=0.112), and *TET2*-CHIP in *BRAF*^mut^ disease was also not significantly associated with PFS (HR 1.86, 95% CI 0.43-8.06, p=0.404).

**Figure 5.**
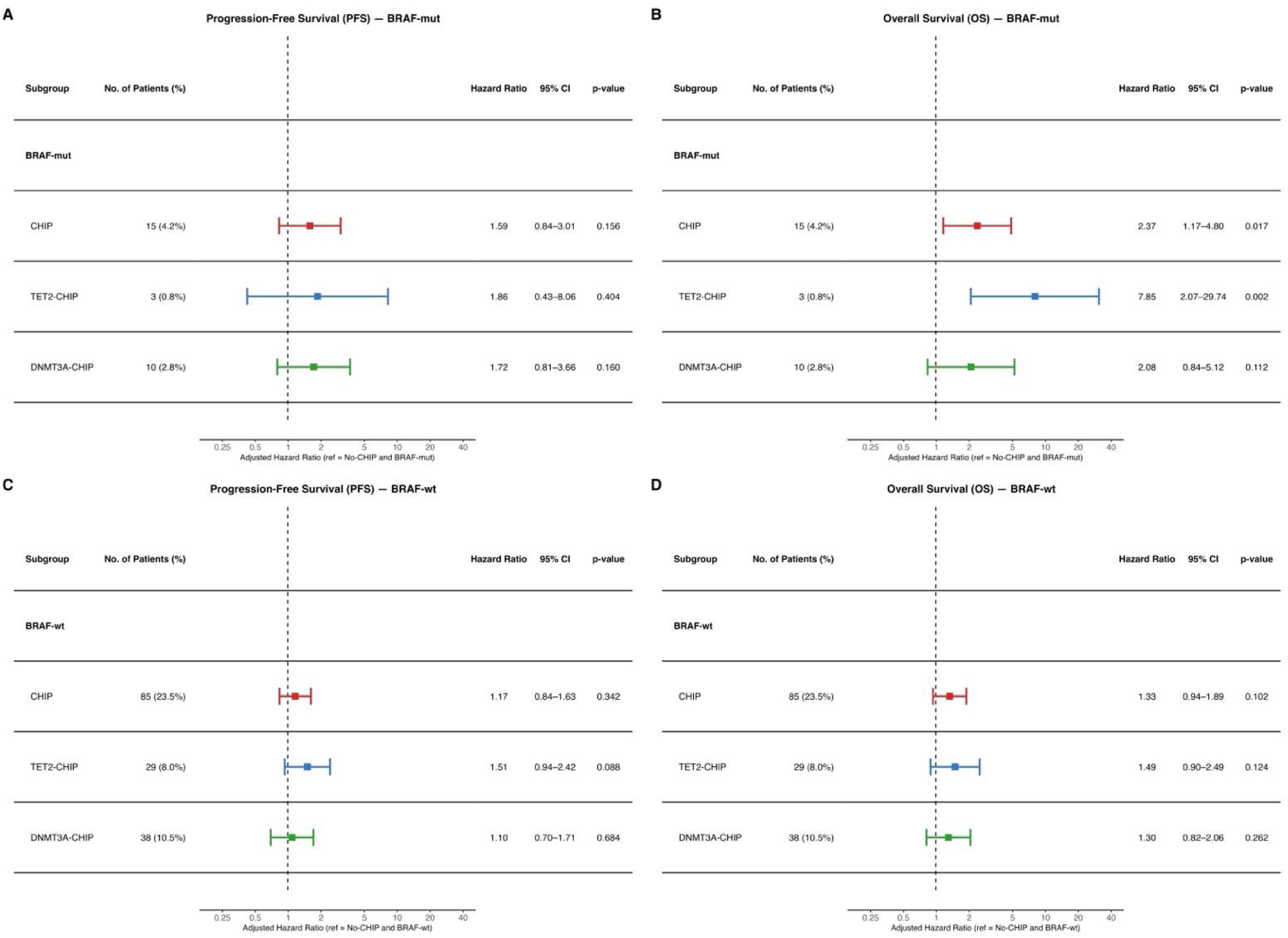
Tabular forest plots of *BRAF*-stratified Cox proportional hazards models evaluating the association of clonal hematopoiesis of indeterminate potential (CHIP) genotype with survival outcomes in 361 patients with unresectable stage III/IV melanoma treated with anti-PD-1–based therapy. Within each *BRAF* parent group, CHIP subgroups include any CHIP, *TET2*-CHIP, and *DNMT3A*-CHIP, with patients without CHIP in the corresponding *BRAF* subgroup serving as the reference group. **(A)** PFS among patients with *BRAF*-mutant melanoma. **(B)** OS among patients with *BRAF*-mutant melanoma. **(C)** PFS among patients with *BRAF*-wild-type melanoma. **(D)** OS among patients with *BRAF*-wild-type melanoma. Squares represent adjusted hazard ratios (HRs) and horizontal whiskers denote 95% confidence intervals (CIs) on a logarithmic scale; the dashed vertical line indicates HR=1.

To formally test whether the association between CHIP and survival differed by tumor *BRAF* genotype, we fit multivariable Cox proportional hazard models including a CHIP × *BRAF* interaction term. Although *BRAF*-stratified estimates suggested more adverse CHIP-associated outcomes among patients with *BRAF*-mutant melanoma, the multiplicative interaction term was not statistically significant for PFS (interaction HR 1.24, 95% CI 0.63–2.46, Wald p=0.538; likelihood ratio test p=0.544) or OS (interaction HR 1.46, 95% CI 0.68–3.10, Wald p=0.329; likelihood ratio test p=0.339). Schoenfeld residual testing did not suggest violation of the proportional hazards assumption for the CHIP × BRAF interaction term. These findings indicate that the BRAF-stratified analyses should be interpreted as exploratory and hypothesis-generating rather than definitive evidence of effect modification by tumor BRAF genotype.

Among patients with *BRAF*^wt^ melanoma, associations between CHIP and outcomes were more modest. CHIP was not significantly associated with PFS (HR 1.17, 95% CI 0.84–1.63, p=0.342) or OS (HR 1.33, 95% CI 0.94–1.89, p=0.102). *TET2*-CHIP in *BRAF*^wt^ disease showed a trend toward worse PFS (HR 1.51, 95% CI 0.94–2.42, p=0.088) but was not significantly associated with OS (HR 1.49, 95% CI 0.90–2.49, p=0.124). *DNMT3A*-CHIP in *BRAF^wt^* disease was not associated with either PFS (HR 1.10, 95% CI 0.70–1.71, p=0.684) or OS (HR 1.30, 95% CI 0.82–2.06, p=0.262).

Taken together, these analyses suggest that CHIP-associated risk may be more adverse among patients with *BRAF^mut^* melanoma, particularly those with *TET2*-CHIP, but formal interaction testing did not establish statistically significant effect modification by tumor *BRAF* genotype.

### Multiple CHIP clones are associated with inferior clinical outcomes

In multivariable Cox models incorporating tumor- and CHIP-related covariates, several factors were associated with inferior outcomes (**Supplementary Figure 4)**. For PFS, *BRAF^mut^* disease was independently associated with worse outcome (HR 1.69, 95% CI 1.23-2.31, p=0.001). CHIP was not independently associated with PFS (HR 1.21, 95% CI 0.90-1.61, p=0.202), nor was CHIP with VAF≥10% (HR 1.19, 95% CI 0.74-1.91, p=0.482). However, concurrent CHIP and *BRAF*^mut^ positivity were associated with significantly worse PFS (HR 2.22, 95% CI 1.25-3.92, p=0.006), and patients with ≥2 CHIP clones at VAF ≥2% had markedly inferior PFS (HR 3.53, 95% CI 1.62-7.72, p=0.002). Age >65 years, stage IV disease, and LDH ≥225 were not significantly associated with PFS in the adjusted model. Because the *BRAF*-stratified analyses and multivariable model use different reference groups and covariate structures, the CHIP/*BRAF*^mut^ estimates should be interpreted as related but non-identical measures of context-dependent risk. This joint-exposure term should not be interpreted as a formal interaction test. For OS, CHIP was independently associated with worse outcome (HR 1.43, 95% CI 1.05-1.93, p=0.023), whereas *BRAF*-mutant status alone was not (HR 1.08, 95% CI 0.75-1.55, p=0.691). CHIP with VAF≥10% was not significantly associated with OS (HR 1.20, 95% CI 0.71-2.02, p=0.503). In contrast, concurrent CHIP and *BRAF*-mut status remained significantly associated with worse OS (HR 2.11, 95% CI 1.13-3.95, p=0.019), as did the presence of ≥2 CHIP clones at VAF ≥2% (HR 3.54, 95% CI 1.62-7.75, p=0.002) and baseline LDH ≥225 (HR 1.52, 95% CI 1.12-2.07, p=0.007). These findings support a model in which CHIP contributes to adverse outcomes in melanoma in a context-dependent manner, with particularly poor outcomes among patients with *BRAF*^mut^ disease and among those with evidence of greater clonal burden.

## DISCUSSION

This study integrates data from human cohorts and a *BRAF-*mutant murine model to support an association between CHIP, particularly *TET2*-driven CHIP, and inferior melanoma outcomes. In a large, treatment-unselected melanoma cohort, CHIP was enriched compared with (1) cancer-free healthy controls (2) non-melanoma cancer controls, and (3) 1:4 age/sex-matched healthy control cohorts, supporting a broader association between CHIP and melanoma beyond the ICI-treated setting. The CHIP enrichment observed in these melanoma cases motivated further analysis in a syngeneic *BRAF*^mut^ melanoma murine model, in which mice with *TET2*-CHIP were found to have significantly increased primary melanoma growth compared with no-CHIP controls. These genotype-specific findings, in conjunction with recent studies highlighting the immunomodulatory properties of CHIP and their associated impact on ICI therapy response, prompted further investigation into a cohort of 361 patients with unresectable stage III/IV melanoma treated with ICI therapy. Within this cohort, CHIP was enriched compared with age/sex-matched healthy controls, and CHIP was associated with significantly worse OS. *TET2*-CHIP showed the clearest adverse association, whereas *DNMT3A*-CHIP did not demonstrate similar risk. In exploratory *BRAF*-stratified analyses, CHIP-positive/*BRAF*-mutant melanoma showed adverse OS estimates, although formal interaction testing and small subgroup sizes limit inference. Associations with CHIP clone size were less consistent, whereas gene-specific analyses suggested that adverse associations were most apparent among patients with *TET2-*CHIP. These findings suggest that the prognostic significance of CHIP may depend in part on tumor *BRAF* genotype.

Our study adds to emerging evidence that genotype-specific CHIP shapes solid tumor outcomes [14, 41, 46]. Consistent with the broader concept that CHIP is biologically heterogeneous, prior retrospective studies have reported that clinical associations vary across CHIP driver genes in several non-melanoma disease contexts [47–49]. Previous groups have also reported *DNMT3A* and *TET2* as common drivers of clonal expansion following ICI therapy, but included patients with both melanoma and NSCLC in the study cohort [29]. By pairing human cohort analyses with syngeneic *BRAF*-mutant melanoma models, our study provides an opportunity to evaluate how distinct CHIP genotypes influence melanoma biology in a controlled experimental setting. This framework also allowed us to examine CHIP-associated tumor growth in the context of *BRAF* mutation, a defining genomic feature present in approximately 40–66% of cutaneous melanomas [50].

Our study has several limitations. First, CHIP and *BRAF* genotype were not evenly distributed in the ICI-treated cohort, which complicates interpretation of their independent and joint effects. CHIP was less common among patients with *BRAF*^mut^ tumors than among those with *BRAF*^wt^ disease (17.6% vs 30.8%), and conversely, *BRAF* mutations were less frequent among patients with CHIP than among those without CHIP (15.0% vs 26.8%). Second, the small size of certain subgroups, including the *TET2*-CHIP/*BRAF*^mut^ subgroup and patients with *TET2*-CHIP or *DNMT3A*-CHIP with VAF ≥10%, limits interpretation of our findings regarding VAF-dependent risk and *BRAF*-stratified survival outcomes. Finally, this study relied on single-timepoint sampling, as whole blood was sequenced only once prior to initiation of anti-PD-1-based therapy. Because CHIP is dynamic, clone size and composition may have evolved over time with treatment in ways that were not captured in this analysis. In addition, because this was a retrospective single-institution study, residual confounding and ascertainment bias remain important limitations. Patients entered the BioVU, BRAF-sequenced, and ICI-treated cohorts through different clinical and sequencing pathways, and BRAF testing and ICI exposure may reflect disease biology, treatment era, clinician selection, or availability of tissue for sequencing. Although multivariable models adjusted for available clinical covariates, unmeasured disease burden, treatment selection, comorbid illness, and CHIP-associated non-melanoma mortality may have influenced OS estimates. Accordingly, BRAF-stratified and interaction analyses should be interpreted as hypothesis-generating.

Despite these limitations, this study expands our understanding of the clinical significance of CHIP in melanoma, suggesting that CHIP genotype, clone size, and tumor BRAF genotype may jointly inform melanoma outcomes, pending validation. Ultimately, this and future work will be critical to determine how somatic hematopoietic mutations should be incorporated into prognostic assessment and treatment selection for patients with solid tumors.

## Data Availability

All data produced in the present study are available upon reasonable request to the authors

## DECLARATIONS

### Ethics approval and consent to participate

The ICI-treated advanced melanoma cohort study was approved by the Vanderbilt University Medical Center Institutional Review Board (IRB #100178). The treatment-unselected melanoma cohort was analyzed using BioVU, the Vanderbilt University Medical Center de-identified biorepository linking electronic health record data to genomic data. All animal procedures were approved by the Vanderbilt University Medical Center Institutional Animal Care and Use Committee and were performed in accordance with institutional and regulatory guidelines.

### Consent for publication

Not applicable. This manuscript does not contain identifiable individual patient data, images, or case details requiring consent for publication.

### Availability of data and material

The clinical and genomic data analyzed in this study are not publicly available because they contain protected or potentially identifiable patient information and are subject to institutional review board and BioVU data-use restrictions. De-identified data or analytic code may be made available from the corresponding author upon reasonable request and subject to applicable institutional approvals, data-use agreements, and regulatory requirements.

### Authors’ contributions

A.K., M.A.H., S.C.R., A.G.B. conceived the study. M.A.H, S.C.R., P.B.F. and A.K. designed the methodology; M.A.H. and S.C.R. performed formal analyses; M.A.H, S.C.R., D.B.J., and A.K. curated clinical data and S.C.R and C.P. performed mouse experiments; M.A.H. and S.C.R. developed the software and visualizations. P.B.F, B.H.P, A.G.B. and A.K. provided resources. M.A.H., S.C.R., and A.K. drafted the manuscript; all authors critically revised it for important intellectual content. A.K., A.G.B., and B.H.P. supervised the project and obtained funding. A.K. and A.G.B. are guarantors and accept full responsibility for the work. All authors approved the final version.

## List of Abbreviations

ASXL1: additional sex combs like 1
ATCC: American Type Culture Collection
BRAF: B-Raf proto-oncogene
BRAFmut: BRAF-mutant
BRAFwt: BRAF-wild-type
CI: confidence interval
CHIP: clonal hematopoiesis of indeterminate potential
DNMT3A: DNA methyltransferase 3 alpha
HR: hazard ratio
IACUC: Institutional Animal Care and Use Committee
ICI: immune checkpoint inhibitor
LDH: lactate dehydrogenase
NSCLC: non-small cell lung cancer
OR: odds ratio
OS: overall survival
PBS: phosphate-buffered saline
PCR: polymerase chain reaction
PD-1: programmed cell death protein 1
PFS: progression-free survival
TET2: ten-eleven translocation 2
VAF: variant allele fraction
VUMC: Vanderbilt University Medical Center

---

**Supplementary Figure 1.**
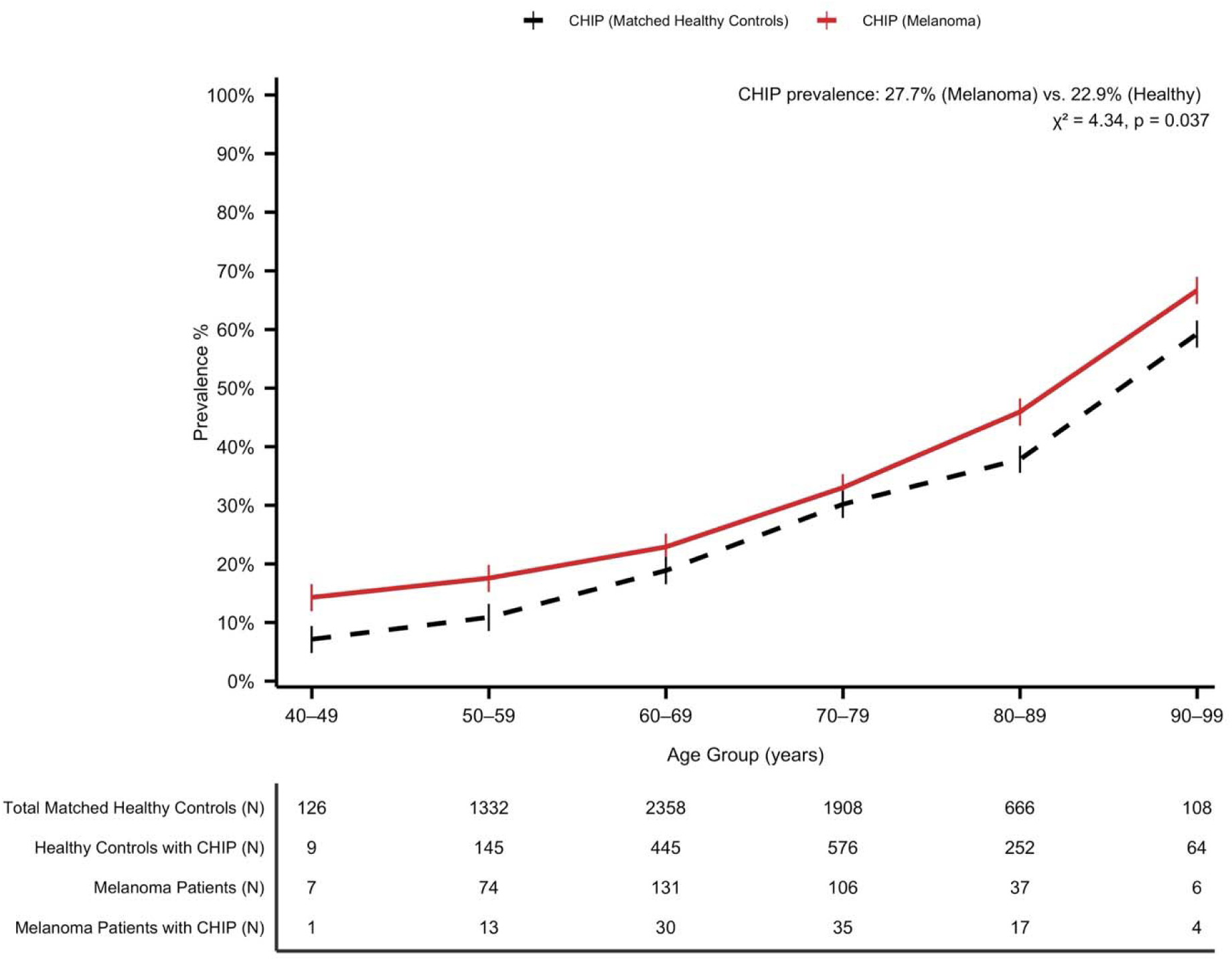
Line plot showing the prevalence of CHIP across 10-year age groups (40–49 through 90–99) in the ICI-treated advanced melanoma cohort (red line) and matched healthy controls (black dashed line). The tabular panel below the plot reports the total number of individuals in each age group and the corresponding number with CHIP in each cohort. Overall CHIP prevalence was higher in melanoma patients than matched healthy controls (27.7% vs 22.9%; χ² = 4.34, p = 0.037).

**Supplementary Figure 2.**
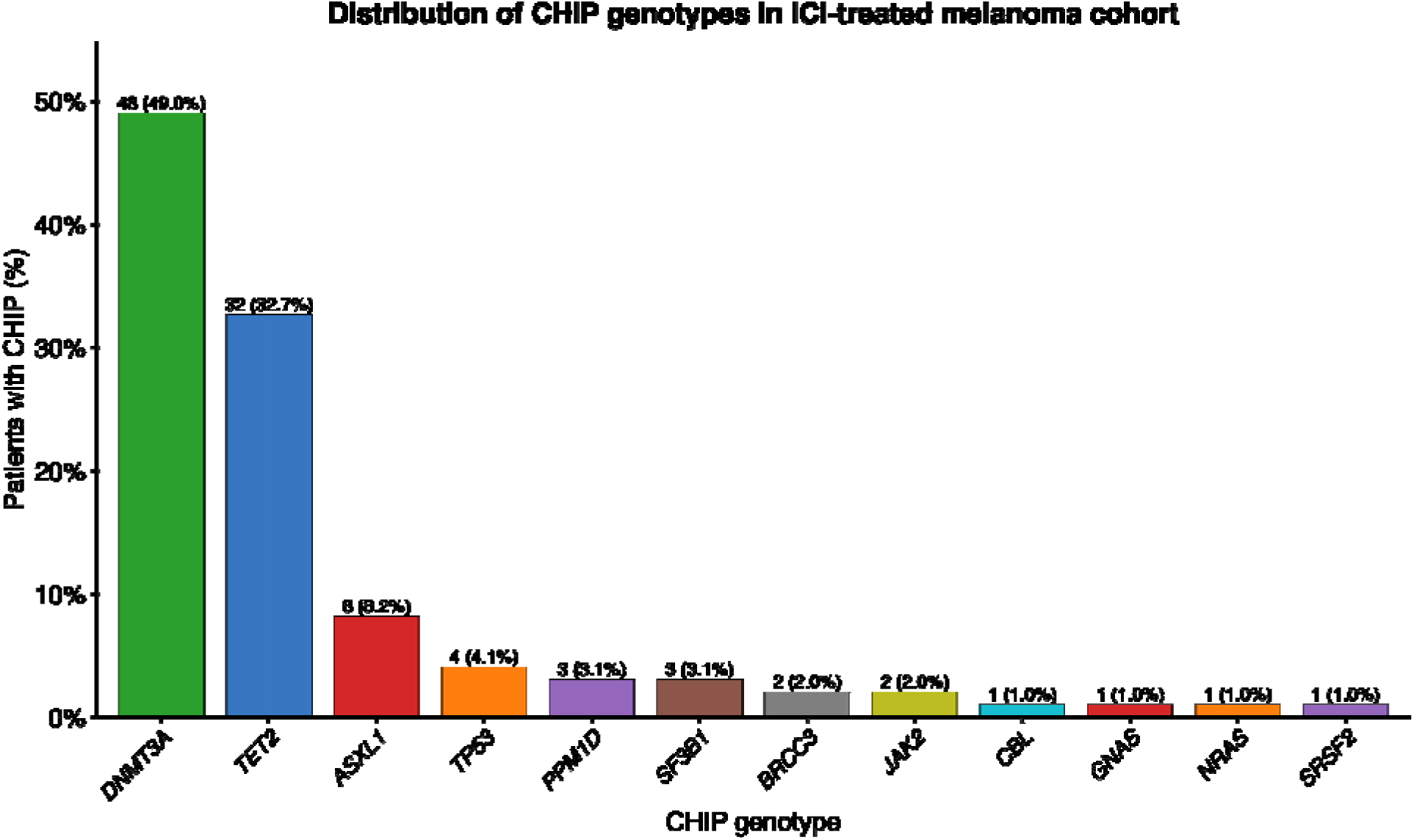
Bar chart showing distribution of CHIP genotypes within the ICI-treatment advanced melanoma cohort. The x-axis displays CHIP-driver genes and the y-axis displays the frequency of each genotype among patients with CHIP. Labels above each bar indicate the absolute number of patients and the corresponding percentage out of patients in the melanoma cohort with CHIP. *DNMT3A* and *TET2* were the most frequently detected CHIP genotypes, followed by *ASXL1* and *TP53*, while other CHIP-associated genotypes were identified at lower frequencies.

**Supplementary Figure 3.**
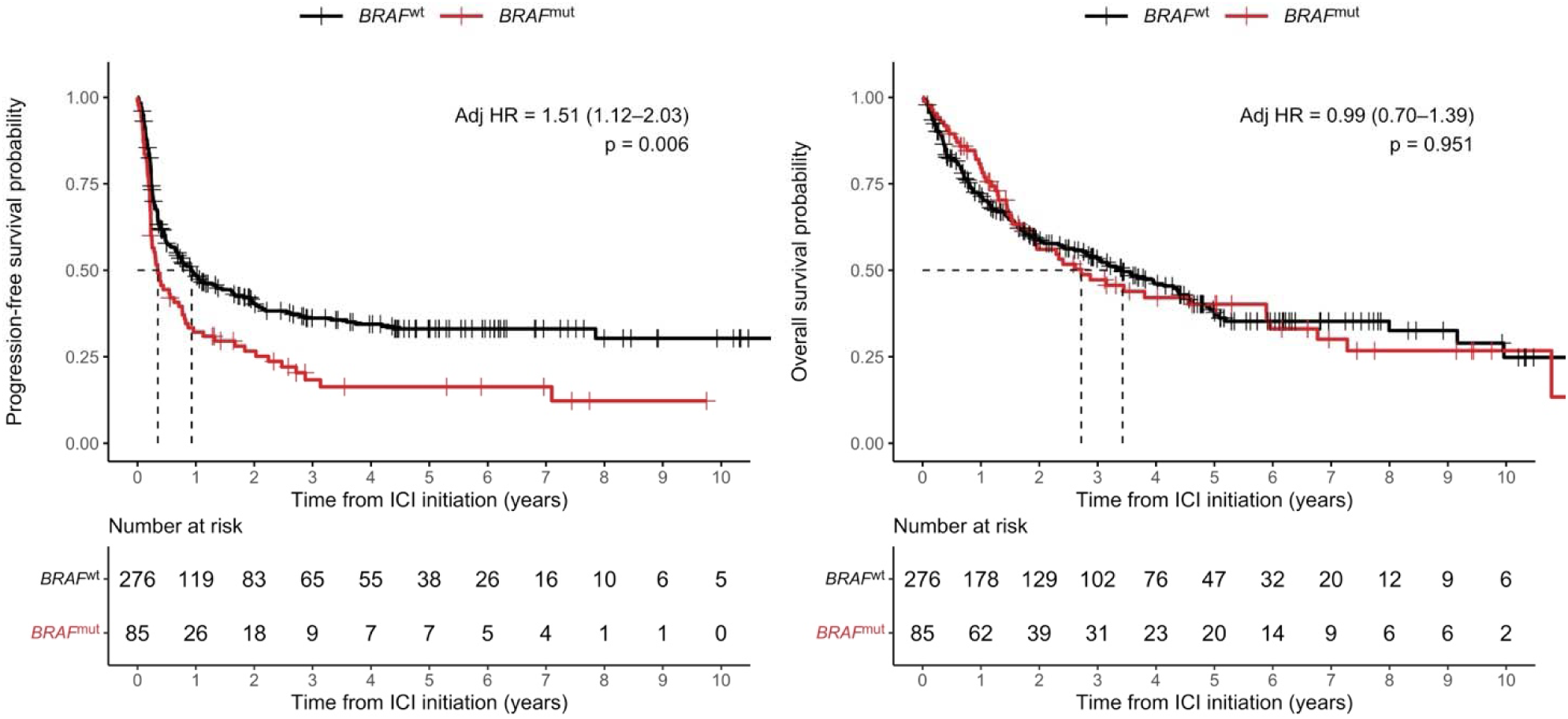
Two-panel Kaplan-Meier plots comparing **(A)** progression-free survival and **(B)** overall survival in years between patients with *BRAF*-mutant versus *BRAF*-wild-type tumors. Adjusted hazard ratios and 95% confidence intervals are from Cox proportional hazards models adjusted for age, sex, stage at anti-PD-1 initiation, and prior therapy before anti-PD-1.

**Supplementary Figure 4.**
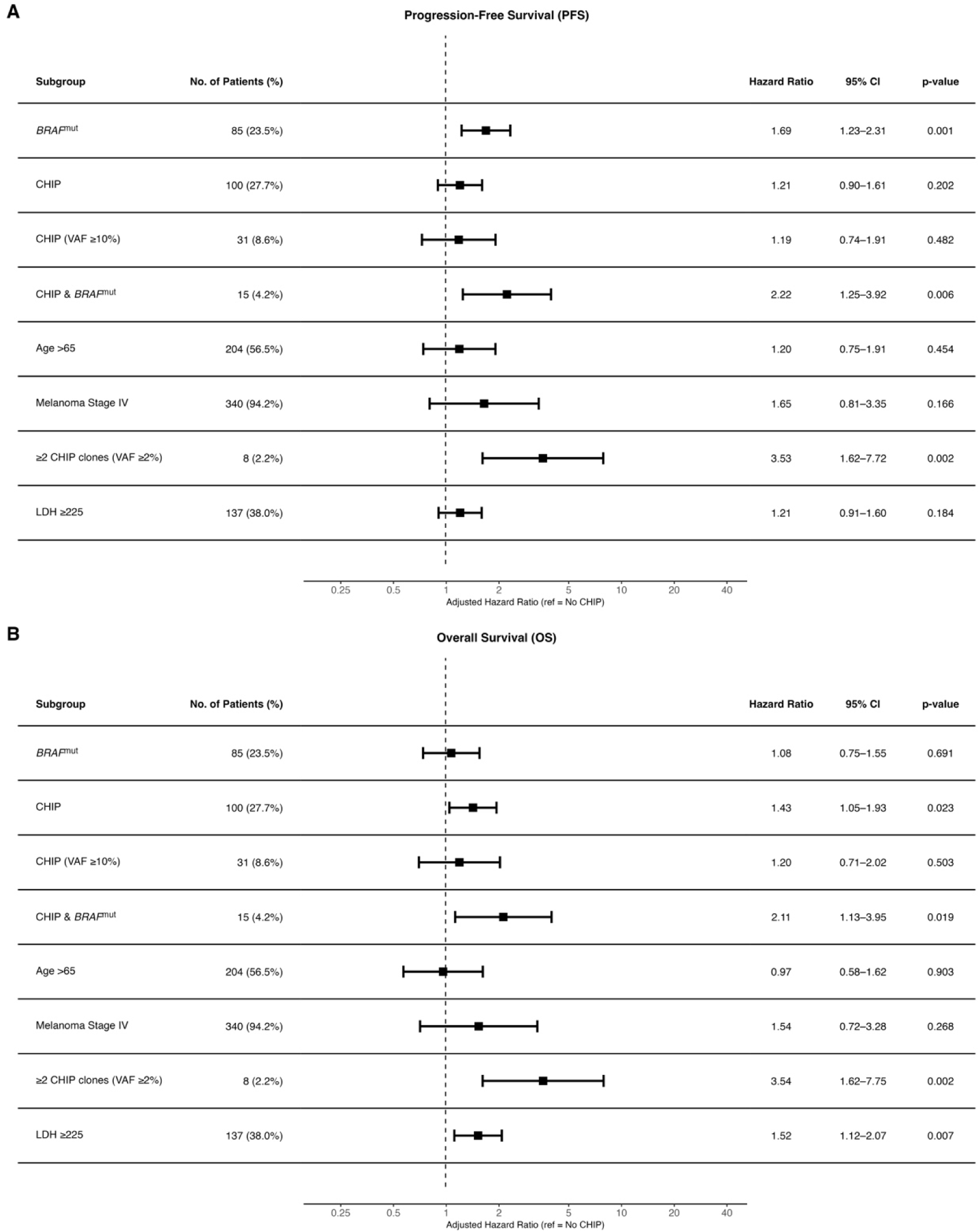
Tabular-forest plot of multivariable Cox proportional hazards models evaluating associations between CHIP and tumor-related factors and survival outcomes in 361 patients with unresectable stage III/IV melanoma treated with anti-PD-1–based therapy. Panel **(A)** shows PFS and Panel **(B)** shows OS. CHIP metrics were derived from whole-blood sequencing using maximum VAF per patient, including CHIP with max VAF ≥10%, ≥2 CHIP clones with VAF ≥2%, and combined CHIP & *BRAF*^mut^ positivity; additional covariates were *BRAF*^mut^ status, age >65 years, any CHIP (VAF≥2%), stage IV disease, and LDH >225 U/L. Squares represent adjusted HRs and horizontal whiskers denote 95% CIs on a logarithmic scale; the dashed vertical line indicates HR=1. The combined CHIP & BRAFmut term represents a joint-exposure estimate, not a formal interaction term.

**Supplementary Table 1.**
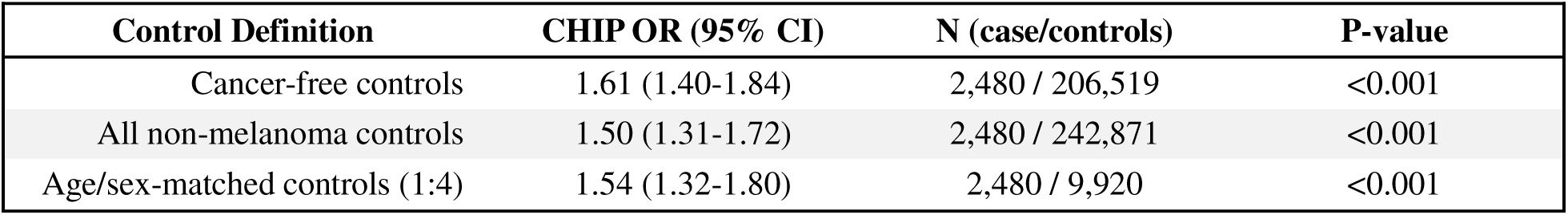
CHIP prevalence in melanoma cases versus controls (adjusted)

| Control Definition | CHIP OR (95% CI) | N (case/controls) | P-value |
| --- | --- | --- | --- |
| Cancer-free controls | 1.61 (1.40-1.84) | 2,480 / 206,519 | <0.001 |
| All non-melanoma controls | 1.50 (1.31-1.72) | 2,480 / 242,871 | <0.001 |
| Age/sex-matched controls (1:4) | 1.54 (1.32-1.80) | 2,480 / 9,920 | <0.001 |

**Supplementary Table 2.**
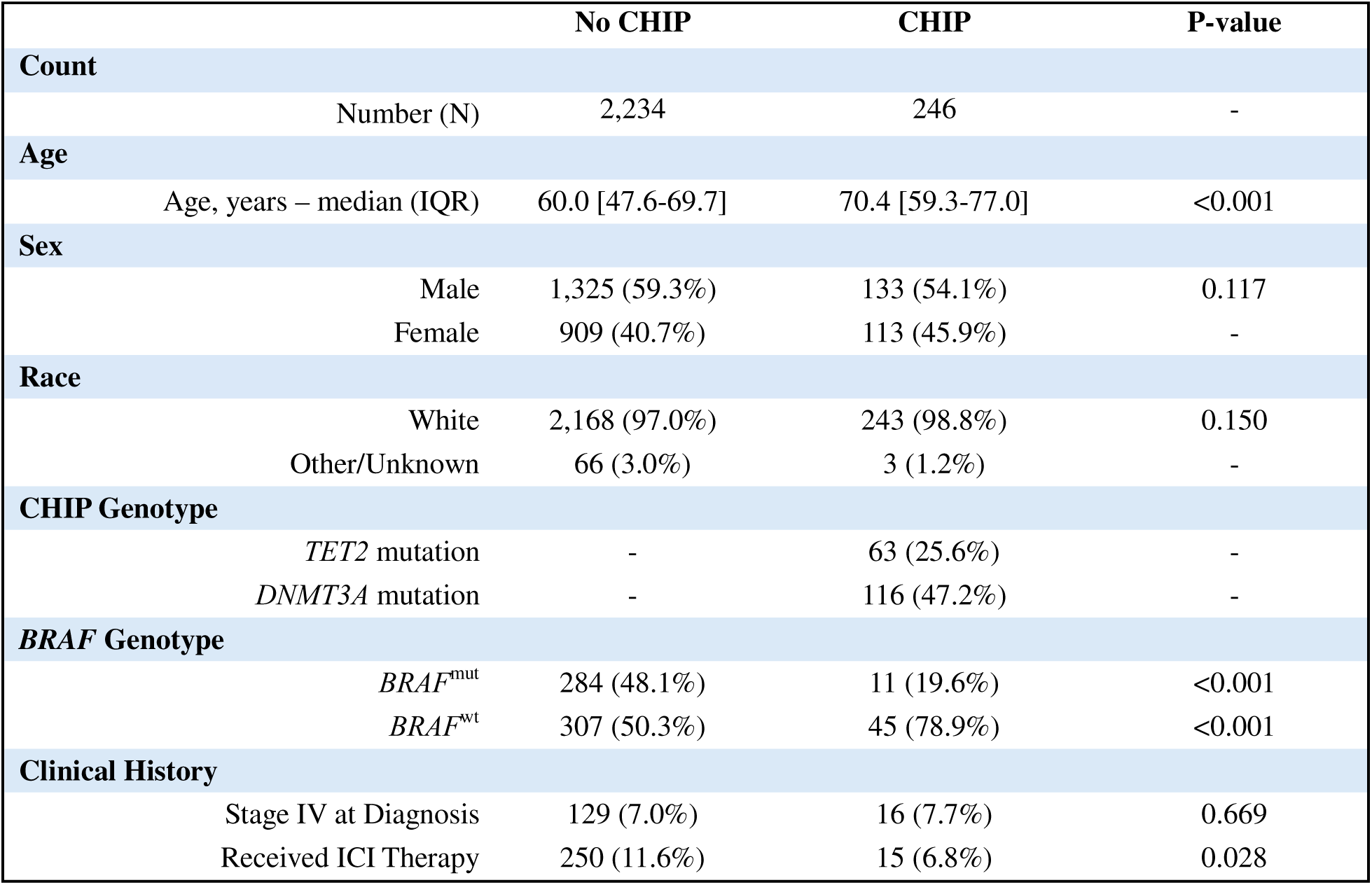
Melanoma cases with overall survival follow-up, by CHIP status.

**Supplementary Table 3.** BioVU melanoma cases with BRAF somatic sequencing, by CHIP status.

|  | No CHIP | CHIP | P-value |
| --- | --- | --- | --- |
| **Count** |  |  |  |
| Number (N) | 591 | 56 | - |
| **Age** |  |  |  |
| Age, years – median (IQR) | 56.9 [44.9-65.9] | 65.6 [55.0-73.1] | <0.001 |
| **Sex** |  |  |  |
| Male | 348 (58.9%) | 35 (62.5%) | 0.670 |
| Female | 243 (41.1%) | 21 (37.5%) | - |
| **Race** |  |  |  |
| White | 578 (97.8%) | 55 (98.2%) | 1.000 |
| Other/Unknown | 13 (2.2%) | 1 (1.8%) | 1.000 |
| **CHIP Genotype** |  |  |  |
| TET2 mutation | - | 16 (28.6%) | - |
| DNMT3A mutation | - | 25 (44.6%) | - |
| **BRAF Genotype** |  |  |  |
| BRAF<sup>mut</sup> | 284 (48.1%) | 11 (19.6%) | <0.001 |
| BRAF<sup>wt</sup> | 307 (51.9%) | 45 (80.4%) | <0.001 |
| **Clinical History** |  |  |  |
| Stage IV at Diagnosis | 69 (15.6%) | 7 (16.7%) | 0.826 |
| Received ICI Therapy | 120 (21.8%) | 6 (12.0%) | 0.145 |

**Supplementary Table 4.** CHIP prevalence in BRAF^mut^ versus BRAF^wt^ melanoma (adjusted)

| CHIP Subgroup | OR, BRAF<sup>mut</sup> vs BRAF<sup>wt</sup> (95% CI) | Number (n) | P-value |
| --- | --- | --- | --- |
| Any CHIP | 0.36 (0.17-0.72) | 56 | 0.006 |
| TET2-CHIP | 0.70 (0.18-2.23) | 16 | 0.562 |
| DNMT3A-CHIP | 0.41 (0.14-1.07) | 25 | 0.084 |

